# Factors associated with nonessential workplace attendance during the Covid-19 pandemic in the UK in early 2021: evidence from cross-sectional surveys

**DOI:** 10.1101/2021.03.30.21254333

**Authors:** Susan Michie, Henry WW Potts, Robert West, Richard Amlot, Louise E Smith, Nicola T Fear, G James Rubin

## Abstract

**Background and aims:** Working from home where possible is important in reducing spread of Covid-19. In early 2021, a quarter of people in England who believed they could work entirely from home reported attending their workplace. To inform interventions to reduce this, this study examined associated factors.

**Methods:** Data from the ongoing CORSAIR survey series of nationally representative samples of people in the UK aged 16+ years in January-February 2021 were used. The study sample was 1422 respondents who reported that they could work completely from home. The outcome measure was self-reported workplace attendance at least once during the preceding week. Factors of interest were analysed in three blocks: 1) sociodemographic variables, 2) variables relating to respondents’ circumstances, and 3) psychological variables.

**Results:** 26.8% (95%CI=24.5%-29.1%) of respondents reported having attended their workplace at least once in the preceding week. Sociodemographic variables and living circumstances significantly independently predicted non-essential workplace attendance: male gender (OR=1.85,95%CI=1.33-2.58), dependent children in the household (OR=1.65,95%CI=1.17-2.32), financial hardship (OR=1.14,95%CI=1.08-1.21), socio-economic grade C2DE (OR=1.74, 95%CI=1.19-2.53), working in sectors such as health or social care (OR=4.18, 95%CI=2.56-6.81), education and childcare (OR=2.45, 95%CI=1.45-4.14) and key public service (OR=3.78, 95%CI=1.83-7.81), and having been vaccinated (OR=2.08,95%CI=1.33-3.24).

**Conclusions:** Non-essential workplace attendance in the UK in early 2021 during the Covid-19 pandemic was significantly independently associated with a range of sociodemographic variables and personal circumstances. Having been vaccinated, financial hardship, socio-economic grade C2DE, having a dependent child at home, working in certain key sectors were associated with higher likelihood of workplace attendance.

## Introduction

Workplaces have been identified as settings for the spread of Covid-19 (EMG Transmission Group, 2021; Mutambudzi et al., 2020), with outbreaks and clusters being reported in a variety of occupational settings in the UK and Europe (European Centre for Disease Prevention and Control, 2020). Factors associated with workplace outbreaks have been found to include occupations associated with low socioeconomic status, workers in essential settings who cannot work from home, and workplaces without robust ‘Covid-19 safe’ policies and procedures (EMG Transmission Group, 2021). Understanding the factors that contribute to people attending the workplace when they do not need to will inform interventions to reduce this practice.

In the UK, national lockdown restrictions were interspersed with regional tiered restrictions to reduce the nature and extent of interpersonal contacts that lead to infectious disease transmission. These restrictions have taken various forms, and have had varied effects, with stringent restrictions shown to outperform more relaxed restrictions in terms of their impact on behaviour, hospitalisations and deaths (Davies et al., 2020). It is therefore likely that contact between people in workplaces contributes to transmission within workplaces and between workplaces and homes (UK Cabinet Office, 2021). Indeed, 40% of people testing positive for Covid-19 reported prior workplace or education activity, and the emergence of clusters have been interpreted to be the result of widespread failure to control risks of airborne and surface transmission in workplaces (HSE and Covid at Work, 2021; Public Health England, 2021).

In many cases, attending the workplace is not essential, either because workers can be furloughed or because people can work from home. However, in the lockdown in early 2021, the third lockdown in England, the Office for National Statistics reported that 48% of working age adults had travelled to work at least once in the past seven days (Office of National Statistics, 2021a). This compared to 37% in the first lockdown in May 2020 (Office of National Statistics, 2020). This may have been associated with more furlough requests having been turned down (TUC, 2021).

Concern has been raised that many people who are attending work at present do not need to do so, and could instead work from home (Strategic Advisory Group on Emergencies, 2021c). The CORSAIR UK national survey of 2000 people found that in February 2021, 35% of those who could work from home had been out to work at least once in the previous week, with 12% at least five times (Department of Health and Social Care, Cabinet Office, 2021). A national poll of nearly 1000 employees commissioned by the TUC and conducted by YouGov found that 19% of those still working were going into offices or other workplaces for part or all of their working week despite being able to work from home (YouGov, 2020b). The main reason given was pressure from employers (40%).

There may be many factors influencing workplace attendance when home working is possible. These may include factors relating to sociodemographic characteristics such as age, gender and ethnic group. For example, young people may perceive themselves to be less at risk from Covid-19 and therefore more likely to attend their workplace. Secondly, they may include factors relating to people’s circumstances, for example the type of job that they do or their living circumstances. For example, people may feel pressure from their employer to attend the workplace or worried about losing their job if they work from home. Or they may be able to work from home but not have adequate equipment to make this easy. And thirdly, they may include factors relating to knowledge and attitudes, for example, being less concerned about the harmfulness of Covid-19 following vaccination.

The roll-out of the vaccination programme in the UK has raised concerns that it may create a sense of reassurance about getting, being harmed by and spreading Covid-19 and that this may lead to more risky behaviours (Strategic Advisory Group on Emergencies, 2021b). This concern arose from evidence of risk compensation and reduced protective behaviours following vaccination from other programmes (Brewer et al., 2007; Reiber et al., 2010), in addition, to the UK survey finding that 29% of respondents said that they would adhere less strictly than before vaccination (YouGov, 2020c) and 22% said they believed that those who had been vaccinated should not be subject to any more coronavirus restrictions (YouGov, 2020a). Real-world data have shown spikes in infection rates in the nine days following vaccination in both Israel (Hunter & Brainard, 2021) and the UK (Bernal et al., 2021), with some suggesting that this may reflect more risky behaviours following vaccination (Rubin et al., 2021).

Understanding factors influencing non-essential workplace attendance during a critical period in the Covid-19 pandemic in the UK could provide useful information to inform interventions aimed at reducing it. This study aimed to examine factors associated with attending the workplace amongst those who could work entirely from home. To do this, we analysed data from the CORSAIR study, designed to collect information during the pandemic to help inform policies and interventions (Smith et al., 2021). This is an ongoing series of surveys carried out weekly or fortnightly with nationally representative samples of UK-based adults. Questions are added in specific waves to address issues of concern at that time.

The variables of interest were analysed according to a model whereby the sociodemographic factors may be expected to have their effect through, and be supplemented by, situational factors which may, in turn, have their effect through and be supplemented by psychological factors. In practice, because it is not possible to measure all the potential predictors of unnecessary workplace attendance with sufficient accuracy, the more distal factors may independently predict attendance, operating through more proximal factors that have not been measured or have not been measured with sufficient precision.

The research question addressed by this study was which variables independently predict nonessential workplace attendance in terms of 1) sociodemographic factors only, 2) sociodemographic factors, personal circumstances and situational factors, and 3) sociodemographic factors, personal circumstances and psychological factors?

## Methods

### Design

The study used data from an ongoing series of cross-sectional online surveys, conducted by BMG Research, a Market Research Society Company Partner, on behalf of the Department of Health and Social Care. The survey began in January 2020 and has continued into 2021 either weekly or fortnightly. Further details are described in Smith et al. (2021). Three waves of the survey were used in which a question about working from home was included: 25-26 January 2021, 8-9 February 2021, and 22-23 February 2021 (waves 42, 43 and 44). Because of the need for rapid turn-around for data collection during a rapidly evolving crisis, the surveys used standard opinion polling methods including nonprobability sampling, an approach common within market research, political polling and social science. Quota samples aim to minimise bias by filling pre-determined targets so that the social and demographic characteristics of the participants match the national population. As such, participants that belong to a quota that has already been met are prevented from completing the survey. Therefore, response rate is not a useful indicator of response bias in quota samples.

## Setting

### United Kingdom

#### Participants

The sample was those who said they could fully work at home, recruited from two specialist online panel providers, Respondi and Savanta (Smith et al., 2021). Participants were eligible for the study if they were aged 16 years or over and lived in the UK. If a respondent completed the survey, they were unable to participate in the following three waves. Quotas were applied based on age and gender (combined) and Government Office Region, and reflected targets based on data from the Office for National Statistics (Office for National Statistics, 2019). Therefore, the socio-demographic characteristics of participants in each survey wave were broadly similar to those in the UK general population. Participants were reimbursed in points which could be redeemed in cash, gift vouchers or charitable donations (up to 70p per survey). The total sample from the three survey waves was 6,033, of whom 3,271 reported that they were in work. Of these 1,422 reported that they could fully work at home and this formed the sample for the present study.

#### Measures

For the outcome measure, participants were asked to “Please enter the number of times you have been out of your home in the last seven days, for each of the following reasons” with “to go out to work” listed as one of the reasons. Responses were dichotomised into any workplace attendance (1) versus none (0). Since the sample has reported that they could work fully at home, we have taken this as a measure of nonessential workplace attendance.

Potential predictor variables were: 1) sociodemographic variables (gender, age, educational level, ethnic group (Office for National Statistics, 2015), English not as first language, Government Office Region in England plus Scotland, Wales and Northern Ireland, survey wave), 2) variables relating to the respondents’ circumstances (marital status, living alone, having a dependent child in the household, employment status, manual occupation of main earner (socioeconomic grade C2DE (National Readership Survey, 2021)), working in one of a number of potentially risky types of workplace, Index of Multiple Deprivation in quartiles (ESRI, 2021), Covid-19-related financial hardship, having a chronic illness that heightens risk of severe illness from Covid-19, having a household member who has a chronic illness, having been vaccinated), and 3) psychological variables (worry about Covid-19, perceived risk to self from Covid-19, believing that one has had Covid-19, belief that Government information on Covid-19 is biased, and willingness to leave the home if they had symptoms). Full details of all the measures are provided in the Supplementary file.

The categories used in the question on work sector were chosen to identify those that work in a key occupational sector. People were asked to, “indicate if you work in any of the following sectors or roles? Please include any voluntary work”. This categorisation may thus not represent a person’s main employment.

#### Ethics

This work was conducted as part of service evaluation of the marketing and communications run by the Department of Health and Social Care, and so did not require ethical approval.

#### Patient and public involvement

Lay members served on the advisory group for the project that developed our prototype survey material; this included three rounds of qualitative testing. Due to the rapid nature of this research, the public was not involved in the further development of the materials during the Covid-19 pandemic.

#### Power

The sample size of approximately 1,400 (depending on the analysis) provided >90% power to detect an odds ratio representing a ‘small’ effect size (f^2^=0.02) with 2-tailed alpha of 0.05 in a multi-variable regression with 24 potential predictor variables entered together.

#### Analysis

The sample was weighted by age, gender and region to match the UK aged 16+ years population. The full weighted sample size was 1,428. Missing values were excluded on an analysis-by-analysis basis leading to smaller sample sizes in some cases. Frequencies and percentages were calculated for prevalence, and multivariable logistic regressions were undertaken to determine associations between the primary outcome and predictor variables.

First all predictor variables in Block 1 (sociodemographic variables) were entered. Then variables from Block 2 (respondents’ circumstances) were added to the model. Then variables from Block 3 (psychological variables) were added.

The analyses plan was not pre-registered and so the analyses should be considered exploratory.

## Results

Of a weighted sample of 1,428, 26.7% (n=382, 95%CI=24.5-29.1) attended the workplace in the preceding seven days. Table 1 shows the characteristics of the sample, overall and by attendance at their workplace.

**Table 1:** Participant characteristics and comparison of those attending and not attending their workplace

| Variable | Attended<br>workplace<sup>1</sup><br>% (n) | Did not attend<br>workplace<sup>1</sup><br>% (n) | All<sup>2,3</sup><br>% (n) |
| --- | --- | --- | --- |
| **Block 1: sociodemographic variables** |  |  |  |
| Wave, % (n) |  |  |  |
| Wave 1 (25-26 January 2021) | 25.7 (132) | 74.3 (382) | 36.0 (514) |
| Wave 2 (8-9 February 2021) | 25.1 (114) | 74.9 (341) | 31.9 (455) |
| Wave 3 (22-23 February 2021) | 29.5 (135) | 70.5 (323) | 32.1 (458) |
| Region, % (n) |  |  |  |
| East Midlands | 25.8 (24) | 74.2 (69) | 6.5 (93) |
| East of England | 22.1 (30) | 77.9 (106) | 9.5 (136) |
| London | 37.2 (108) | 62.8 (182) | 20.3 (290) |
| North East | 29.4 (15) | 70.6 (36) | 3.6 (51) |
| North West | 28.4 (42) | 71.6 (106) | 10.4 (148) |
| Northern Ireland | 26.5 (9) | 73.5 (25) | 2.4 (34) |
| Scotland | 20.6 (27) | 79.4 (104) | 9.2 (131) |
| South East | 15.5 (26) | 84.5 (142) | 11.8 (168) |
| South West | 21.7 (20) | 78.3 (72) | 6.4 (92) |
| Wales | 26.8 (15) | 73.2 (41) | 3.9 (56) |
| West Midlands | 28.8 (38) | 71.2 (94) | 9.2 (132) |
| Yorks & Humber | 27.6 (27) | 72.4 (71) | 6.9 (98) |
| Gender, % (n) * |  |  |  |
| Female | 19.8 (125) | 80.2 (505) | 44.2 (630) |
| Male | 32.2 (256) | 67.8 (538) | 55.8 (794) |
| Age, mean (SD) * | ? | ? | ? |
| Educational level, % (n) |  |  |  |
| Non-degree level | 26.5 (177) | 73.5 (492) | 46.8 (669) |
| Degree level | 27.0 (205) | 73.0 (554) | 53.2 (759) |
| Ethnic group, % (n) * |  |  |  |
| White British | 24.0 (271) | 76.0 (856) | 79.2 (1127) |
| White ethnic minorities | 35.2 (37) | 64.8 (68) | 7.4 (105) |
| Ethnic minorities (excluding<br>White minorities) | 38.2 (73) | 61.8 (118) | 13.4 (191) |
| Language, % (n) |  |  |  |
| English as 1<sup>st</sup> language | 25.8 (327) | 74.2 (941) | 88.7 (1268) |
| English not as 1<sup>st</sup> language | 34.2 (55) | 65.8 (106) | 11.3 (161) |
| **Block 2: Personal circumstances** |  |  |  |
| Marital status, % (n) |  |  |  |
| Not married/partnered | 24.9 (120) | 75.1 (362) | 33.9(482) |
| Married/partnered | 27.4 (258) | 72.6 (682) | 66.1 (940) |
| Living social status, % (n) |  |  |  |
| Live with other(s) | 27.3 (327) | 72.8 (873) | 84.0 (1200) |
| Live alone | 24.0 (55) | 76.0 (174) | 16.0 (229) |
| Children at home, % (n) * |  |  |  |
| No dependent children | 18.2 (140) | 81.8 (628) | 53.8 (768) |
| Dependent children | 36.7 (242) | 63.3 (418) | 46.2 (660) |
| Employment status, % (n) * |  |  |  |
| Full time | 29.0 (339) | 71.0 (828) | 81.7 (1167) |
| Part time | 15.5 (24) | 84.5 (131) | 10.9 (155) |
| Self-employed | 17.9 (19) | 82.1 (87) | 7.4 (106) |
| Socio-economic grade, % (n) * |  |  |  |
| Non-manual occupation | 21.7 (234) | 78.3 (846) | 76.5 (1080) |
| Manual occupation | 43.4 (144) | 56.6 (188) | 23.5 (332) |
| Work sector, % (n) * |  |  |  |
| Other | 15.4 (103) | 84.6 (564) | 46.7 (667) |
| Health & social care | 53.0 (105) | 47.0 (93) | 13.9 (198) |
| Education & childcare | 33.6 (45) | 66.4 (89) | 9.4 (134) |
| Key public service | 43.3 (26) | 56.7 (34) | 4.2 (60) |
| Local & national govt | 18.6 (18) | 81.4 (79) | 6.8 (97) |
| Food & essential goods | 37.0 (20) | 63.0 (34) | 3.8 (54) |
| Public safety & security | 44.0 (11) | 56.0 (14) | 1.7 (25) |
| Transport | 38.1 (16) | 61.9 (26) | 2.9 (42) |
| Utilities, comms & finance | 25.0 (38) | 75.0 (114) | 10.6 (152) |
| Index of multiple deprivation, % (n) |  |  |  |
| 1<sup>st</sup> quartile (lowest) | 21.1 (62) | 78.9 (232) | 20.6 (294) |
| 2<sup>nd</sup> quartile | 23.1 (83) | 76.9 (277) | 25.2 (360) |
| 3<sup>rd</sup> quartile | 24.2 (93) | 75.8 (291) | 26.9 (384) |
| 4<sup>th</sup> quartile (highest) | 37.1 (145) | 62.9 (246) | 27.4 (391) |
| Financial hardship, mean (SD, n) | 9.50 (3.01, 371) | 7.75 (2.92, 1016) | 8.21 (3.05, 1387) |
| Chronic illness of self, % (n) |  |  |  |
| No chronic illness | 25.5 (306) | 74.5 (894) | 85.5 (1200) |
| Chronic illness | 34.5 (70) | 65.5 (133) | 14.5 (203) |
| Chronic illness of household member, % (n) |  |  |  |
| No chronic illness | 26.8 (330) | 73.2 (903) | 87.9 (1233) |
| Chronic illness | 27.1 (46) | 72.9 (124) | 12.1 (170) |
| Vaccination status, % (n) * |  |  |  |
| Not been vaccinated | 23.1 (277) | 76.9 (921) | 84.0 (1198) |
| Been vaccinated | 45.4 (104) | 54.6 (125) | 16.0 (229) |
| **Block 3: Psychological variables** |  |  |  |
| Worried about Covid, mean (SD, n) | 2.35 (1.14, 382) | 2.46 (1.10, 1046) | 2.43 (1.11, 1428) |
| Risk of Covid to self, mean (SD, n) | 3.23 (1.20, 380) | 3.15 (1.10, 1034) | 3.19 (1.13, 1414) |
| Believe Gov info is biased, mean (SD, n) | 2.62 (1.14, 382) | 2.92 (1.23, 1046) | 2.84 (1.21, 1428) |
| * |  |  |  |
| Believe had Covid, % (n) * |  |  |  |
| Not had Covid or Don't know | 24.0 (264) | 76.0 (836) | 77.0 (1100) |
| Had Covid | 36.0 (118) | 64.0 (210) | 23.0 (328) |
| Willing to leave home if with symptoms, % (n) * |  |  |  |
| Not willing | 19.1 (160) | 80.9 (677) | 65.2 (837) |
| Willing | 30.4 (136) | 69.6 (311) | 34.8 (447) |

<sup>1</sup>Percentages sum to 100 across 'Attended workplace' and 'Did not attend workplace'. <sup>2</sup>Percentages sum to 100 down level of predictor variable. <sup>3</sup>Sample sizes vary because of missing values and rounding of weighted totals. \* Workplace attendance differs across values of predictor variable $p < 0.001$ by $\chi^2$ -test for percentages or analysis of variance for means.

Table 2 shows the results of the multivariable logistic regression analysis in the three blocks. In the first block, age, gender, ethnic group and educational level were predictive of workplace attendance. With the addition of the second block, age and educational level were no longer significant predictors of workplace attendance while having dependent children at home, socio-economic grade C2DE, financial hardship, having been vaccinated, and working in a certain sectors (health and social care, education and childcare, key public services, food and essential goods, public safety and security and transport) were associated with higher likelihood of workplace attendance. Working part time or being self-employed were associated with lower likelihood of workplace attendance. None of the psychological variables included were significantly associated with workplace attendance.

**Table 2:** Factors associated with attending the workplace at least once in the previous 7 days using multivariable logistic regression^1^

| Predictor | Block 1<br>(Odds ratio,<br>95%CI) | Block 2<br>(Odds ratio,<br>95%CI) | Block 3<br>(Odds ratio,<br>95%CI) |
| --- | --- | --- | --- |
| **Block 1: sociodemographic variables** |  |  |  |
| Wave |  |  |  |
| Wave 1 (Ref) |  |  |  |
| Wave 2 | 0.95, 0.66-1.35 | 0.89, 0.61-1.31 | 0.90, 0.61-1.33 |
| Wave 3 | 1.35, 0.97-1.88 | 1.13, 0.78-1.63 | 1.18, 0.81-1.71 |
| Region |  |  |  |
| East Midlands (Ref) |  |  |  |
| East of England | 0.69, 0.33-1.45 | 0.62, 0.28-1.41 | 0.61, 0.27-1.39 |
| London | 1.10, 0.58-2.09 | 0.96, 0.48-1.94 | 0.95, 0.47-1.93 |
| North East | 1.25, 0.53-2.95 | 1.24, 0.49-3.18 | 1.28, 0.50-3.29 |
| North West | 1.14, 0.57-2.28 | 1.04, 0.49-2.23 | 1.02, 0.47-2.22 |
| Northern Ireland | 1.37, 0.51-3.69 | 1.09, 0.37-3.25 | 1.01, 0.33-3.05 |
| Scotland | 0.71, 0.34-1.47 | 0.72, 0.32-1.58 | 0.71, 0.32-1.57 |
| South East | 0.58, 0.29-1.19 | 0.56, 0.25-1.23 | 0.55, 0.25-1.22 |
| South West | 0.95, 0.44-2.02 | 0.73, 0.31-1.71 | 0.74, 0.31-1.75 |
| Wales | 1.22, 0.52-2.86 | 1.29, 0.51-3.27 | 1.32, 0.52-3.35 |
| West Midlands | 0.96, 0.48-1.93 | 0.77, 0.36-1.67 | 0.74, 0.34-1.61 |
| Yorks & Humber | 1.02, 0.47-2.19 | 0.96, 0.41-2.22 | 0.93-0.40-2.19 |
| Gender |  |  |  |
| Male (Ref) |  |  |  |
| Female | **1.89, 1.42-2.53** | **1.83, 1.32-2.54** | **1.85, 1.33-2.58** |
| Age | **0.97, 0.96-0.99** | 0.99, 0.98-1.01 | 0.99, 0.98-1.01 |
| Educational level |  |  |  |
| Non-degree level (Ref) |  |  |  |
| Degree level | **0.70, 0.52-0.94** | 0.78, 0.56-1.09 | 0.76, 0.54-1.07 |
| Ethnic group |  |  |  |
| White British (Ref) |  |  |  |
| White ethnic minorities | 1.75, 0.91-3.35 | **2.36, 1.15-4.83** | **2.39, 1.16-4.91** |
| Ethnic minorities (excluding White minorities) | **1.68, 1.09-2.60** | 1.49, 0.92-2.41 | 1.49, 0.92-2.42 |
| Language |  |  |  |
| English as 1<sup>st</sup> language (Ref) |  |  |  |
| English not as 1<sup>st</sup> language | 0.78, 0.45-1.37 | 0.66, 0.36-1.24 | 0.69, 0.37-1.29 |
| **Block 2: Personal circumstances** |  |  |  |
| Marital status | - |  |  |
| Not married/partnered (Ref) |  |  |  |
| Married/partnered |  | 1.20, 0.79-1.83 | 1.20, 0.79-1.83 |
| Living social status | - |  |  |
| Live with other(s) (Ref) |  |  |  |
| Live alone |  | 0.83, 0.48-1.43 | 0.82, 0.47-1.43 |
| Dependent children | - |  |  |
| No dependent children (Ref) |  |  |  |
| Dependent children |  | **1.60, 1.14-2.24** | **1.65, 1.17-2.32** |
| Employment status | - |  |  |
| Full time (ref) |  |  |  |
| Part time |  | **0.40, 0.21-0.73** | **0.38, 0.20-0.70** |
| Self-employed |  | **0.43, 0.21-0.87** | **0.43, 0.21-0.89** |
| Socio-economic grade |  |  |  |
| Non-manual occupation (Ref) |  |  |  |
| Manual occupation |  | **1.75, 1.20-2.54** | **1.74, 1.19-2.53** |
| Work sector | - |  |  |
| Other (Ref) |  |  |  |
| Health & social care |  | **4.26, 2.63-6.90** | **4.18, 2.56-6.81** |
| Education & childcare |  | **2.35, 1.40-3.94** | **2.45, 1.45-4.14** |
| Key public service |  | **3.50, 1.70-7.20** | **3.78, 1.83-7.81** |
| Local & national govt |  | 1.06, 0.51-2.22 | 1.07, 0.51-2.23 |
| Food & essential goods |  | **2.56, 1.20-5.48** | **2.51, 1.17-5.37** |
| Public safety & security |  | **3.60, 1.29-10.1** | **3.90, 1.39-11.0** |
| Transport |  | **2.60, 1.18-5.72** | **2.60, 1.17-5.77** |
| Utilities, comms & finance |  | 1.48, 0.89-2.45 | 1.45, 0.87-2.42 |
| Index of multiple deprivation | - |  |  |
| 1<sup>st</sup> quartile (Ref) |  |  |  |
| 2<sup>nd</sup> quartile |  | 0.97, 0.61-1.54 | 0.98, 0.62-1.56 |
| 3<sup>rd</sup> quartile |  | 0.98, 0.62-1.56 | 0.99, 0.63-1.58 |
| 4<sup>th</sup> quartile |  | 1.02, 0.63-1.65 | 1.10, 0.62-1.64 |
| Covid-19-related financial hardship (15-point score) | - | **1.13, 1.07-1.20** | **1.14, 1.08-1.21** |
| Chronic illness of self | - |  |  |
| No chronic illness (Ref) |  |  |  |
| Chronic illness |  | 0.61, 0.37-1.02 | 0.62, 0.37-1.02 |
| Chronic illness of household member | - |  |  |
| No chronic illness (Ref) |  |  |  |
| Chronic illness |  | 1.15, 0.70-1.86 | 1.14, 0.70-1.86 |
| Vaccination status |  |  |  |
| Not been vaccinated (Ref) |  |  |  |
| Been vaccinated |  | **2.01, 1.29-3.11** | **2.08, 1.33-3.24** |
| **Block 3: Psychological variables** |  |  |  |
| Worry about Covid (5-point scale) | - | - | 1.08, 0.91-1.28 |
| Risk of Covid to self (5-point scale) | - | - | 1.12, 0.94-1.32 |
| Believe had Covid | - |  |  |
| Not had Covid (Ref) |  |  |  |
| Had Covid |  |  | 0.66, 0.44-1.00 |
| Believe Govt info is biased (5-point scale) | - | - | 0.96, 0.84-1.10 |
| Willing to leave home if with symptoms | - | - |  |
| Not willing (Ref) |  |  |  |
| Willing |  |  | 1.05, 0.75-1.48 |

<sup>1</sup>Weighted sample size for the analysis was 1,194.

## Discussion

A substantial percentage of the UK population were attending their workplace in early 2021 even though they reported being able to work fully from home, contrary to UK Government guidance (UK Government, 2021). This suggests that there may be scope for reducing transmission by reducing the prevalence of this behaviour. Having been vaccinated, financial hardship, socio-economic grade C2DE, having a dependent child at home, and working in certain key sectors were associated with higher likelihood of workplace attendance. Women were less likely to attend the workplace than men. Several of the predictors of workplace attendance could reflect targets for interventions aimed at reducing Covid-19 transmission.

The association between financial hardship and workplace attendance could mean that people who are struggling to meet their living costs feel greater pressure to attend than others. This may reflect a more precarious working environment and there is evidence of employer pressure playing a role. The TUC has said that people who could work from home should not be pressured to attend workplaces, nor should they be given the option of doing so voluntarily (TUC, 2021). Increasing job security and reducing employer pressure could be addressed by Government regulation. Greater financial and practical support for those asked to isolate could reduce those going out to work: findings from the CORSAIR study show that more than half of those even with symptoms are not isolating for the full period, and going out to work is one of the reasons given (Smith et al., 2021) In terms of the association with the presence of a dependent child in the household, it is notable that this exists even after adjusting for multiple financial variables within the dataset. We note that the mental health of parents was affected by the first lockdown in the UK (Pierce et al., 2020): one explanation may be that some parents attend work partly in order to reduce distress within the household. Another may be that although they can work at home when children are at school, they cannot do so easily when children are at home.

In April 2020, believing you had already had Covid-19 was shown to be associated with perceptions of immunity against the virus and reduced adherence to several protective behaviours in one UK sample (Smith et al., 2020). While we did not observe an association between perceptions of prior infection and attending work, our findings of a positive association between reports of having been vaccinated and workplace attendance suggests that perceptions of immunity arising from the vaccine are now playing a similar role. Although the roll-out of the vaccination programme in the UK has been rapid and is widely considered a success, there have been reported gaps in the provision of verbal and accessible written information explaining that immunity would take three weeks to build up, would be partial and it was possible that people could continue to be infectious, especially before the second dose. The absence of this information may be associated with a recent increase in self-reported breaches of current lockdown restrictions amongst older adults who were among the first to be vaccinated, where 41% of over 80s reported having met someone outside of their household and support bubble less than 3 weeks after vaccination (Office of National Statistics, 2021b). A month or so into the programme, NHS England have provided scripts, posters and an animation for use alongside the vaccination programme (NHS England, 2021) Hopefully, this will go some way to reducing the increased risky behaviours that can follow vaccination.

Increased workplace attendance in certain sectors suggests that these sectors may be considered for targeted interventions. In the case of health and social care, education and childcare and some other sectors, it could be that the fact that many front-line workers in these sectors need to attend the workplace leads to a culture in which other workers feel compelled to do so even if this is not necessary, something that may also have wider implications for attendance while sick (Webster et al., 2019). Personal communication suggests that another key reason for health and social care employees who could work at home not to do so is the lack of adequate digital technology for their work. This merits further examination.

In the UK, the Government’s roadmap out of our third lockdown specifies a sequence of changes, starting with the reopening of schools and progressing to the removal of all legal limits on social contact. The need to proceed slowly through these changes has been emphasised repeatedly. Ensuring that the large number of people who can work from home do so is one key measure that can be taken to keep control of the pandemic as restrictions are eased. Given the importance of schools remaining open and the predicted increased transmission from children being at school (Strategic Advisory Group on Emergencies, 2021a) it is imperative that other measures are taken to keep Covid-19 under control. Providing the large number of people with the means to work at home when this is possible is one such measure, especially since this would reduce people interacting both in workplaces and on transport.

The limitations of our study include: 1) it relies on self-report which may be reporting bias, particularly concerning whether work can be completed entirely from home - it is possible that whilst our respondents reported that they believed they could work from home, we do not know the circumstances of their employment to know whether this reflects the requirements of their role, 2) use of an online sample which, even though it has been weighted to match major demographic features of the UK population, may nevertheless not be fully representative, 3) collinearity among predictors, and 4) possible omission of other relevant variables.

## Conclusions

Nonessential workplace attendance in the UK in early 2021 during the Covid-19 pandemic was substantial and significantly independently associated with a wide range of sociodemographic variables and personal circumstances. Having been vaccinated, financial hardship, manual occupational group, having a dependent child at home, and working in certain key sectors were associated with higher likelihood of workplace attendance. These findings could inform Government, employer-led and other interventions aimed at reducing nonessential workplace attendance in the future.

## Supporting information

Questionnaire items

## Data Availability

The data are owned by the UK Department of Health and Social Care so no additional data are available from the authors.

## Notes

### Competing Interest Statement

Authors had financial support from the UK National Institute for Health Research for the submitted work. RW has undertaken research and consultancy for companies that manufacture smoking cessation medications (Pfizer, GSK). RA is an employee of Public Health England. HWWP receives additional salary support from Public Health England and NHS England. NTF is a participant of an independent group advising NHS Digital on the release of patient data. All authors are members of the UK Scientific Advisory Group for Emergencies or its subgroups. There are no other financial relationships with any organisations that might have an interest in the submitted work in the previous three years and no other relationships or activities that could appear to have influenced the submitted work.

### Funding Statement

LS, RA, and GJR are supported by the National Institute for Health Research Health Protection Research Unit (NIHR HPRU) in Emergency Preparedness and Response, a partnership between Public Health England (PHE), Kings College London, and the University of East Anglia. RA is also supported by the NIHR HPRU in Behavioural Science and Evaluation, a partnership between PHE and the University of Bristol. HWWP receives funding from PHE and NHS England. NTF is part funded by a grant from the UK Ministry of Defence. The views expressed are those of the authors and not necessarily those of the NIHR, PHE, the Department of Health and Social Care, or the Ministry of Defence. Surveys were commissioned and funded by Department of Health and Social Care (DHSC), with the authors providing advice on the question design and selection. DHSC had no role in analysis, decision to publish, or preparation of the manuscript. Preliminary results were made available to DHSC and the UK Scientific Advisory Group for Emergencies.

### Author Declarations

This work was conducted as part of service evaluation of the marketing and communications run by the Department of Health and Social Care, and so did not require ethical approval. We sought advice from the Psychiatry, Nursing and Midwifery Research Ethics Office, King's College London and confirmed with them that this was service evaluation rather than research.

