## Supplementary material for "Factors associated with nonessential workplace attendance during the Covid-19 pandemic in the UK in early 2021: evidence from cross-sectional surveys": Questionnaire items

This file contains details of the questionnaire items used to measure the variables used in the study:

### 1. Region

OPEN RESPONSE

**Postcode. Could you please provide your full UK postcode? Please ensure to include a space where applicable, e.g. AB1 2CD**

### 2. Gender

SINGLE RESPONSE

**Gender. What is your gender?**

| Male | 1 |
| --- | --- |
| Female | 2 |
| Prefer to self-describe | 95 |
| Prefer not to say | 97 |

### 3. Age

NUMBER RESPONSE

**AgeInt. Please can you tell me your age at your last birthday?**

### 4. Educational level

MULTI RESPONSE

**D8. What is the highest level of educational qualification you have received?**

SINGLE CODE

| PHD/Doctor | 1 |
| --- | --- |
| Masters | 2 |
| Bachelor’s Degree or equivalent (Such as a NVQ level 5) | 3 |
| Higher education (Such as a HND or a NVQ level 4) | 4 |
| A level or equivalent (Such as Scottish Highers or NVQ level 3) | 5 |
| GCSE and below (Such as O level or an RSA Diploma) | 6 |
| Other qualifications (Such as NVQ level 1) | 7 |
| No qualifications | 8 |

### 5. Ethnic group

**D7. Which of the following categories would best describe your ethnicity?**

SINGLE CODE

| **White** |  |
| --- | --- |
| British/English/Welsh/Scottish/Northern Irish | 1 |
| Irish | 2 |
| Gypsy, Traveller or Irish Traveller | 3 |
| Any other White background | 4 |
| **Mixed/ Multiple ethnic groups** |  |
| White and Black Caribbean | 5 |
| White and Black African | 6 |
| White and Asian | 7 |
| Any other Mixed/ Multiple ethnic background | 8 |
| **Asian or Asian British** |  |
| Indian | 9 |
| Pakistani | 10 |
| Bangladeshi | 11 |
| Chinese | 12 |
| Any other Asian background | 13 |
| **Black or Black British** |  |
| African | 14 |
| Caribbean | 15 |
| Any other Black/ African/ Caribbean background | 16 |
| **Other ethnic group** |  |
| Arab | 17 |
| Other | 18 |
| Don’t know | 98 |
| Prefer not to say | 99 |

### 6. Language

**D7A. What is your first language?**

Please type in and select from the options below.

(List of languages. Code 0 for English and 1 for all other languages)

### 7. Marital status

**D1D. What is your marital status?**

| Single, never married | 1 |
| --- | --- |
| Married | 2 |
| Separated | 3 |
| Divorced | 4 |
| Widowed | 5 |
| Partnered/in a relationship | 6 |
| Prefer not to say | 7 |

(Code 1 if selected 1 or 6, and 0 otherwise)

### 8. Living social status

**Q19. How many people currently live in your household? Please select one answer**

*Please include yourself and all adults and children – including those not related to you*

1. I live alone
2. 2
3. 3 – 4
4. 5 – 6
5. 7 +

(Code 1 if selected 1 and 0 otherwise)

### 9. Children at home

**D1.** **What age are any dependent children in your household? Please select all that apply**

*Dependent children are those aged under 18 living in your household.*
MULTI CODE

| No dependent children in household | 1 |
| --- | --- |
| 0 – 4 years old | 2 |
| 5 – 10 years old | 3 |
| 11 – 15 years old | 4 |
| 16 – 18 years old | 5 |

(Code 1 if selected 2 to 5 and 0 if selected 1)

### 10. Employment status

**D2**. SINGLE RESPONSE

**What is your employment status**?

| Full time paid job (31+ hours) | 1 |
| --- | --- |
| Part time paid job (<31 hours) | 2 |
| Doing paid work on a self-employed basis or within your own business | 3 |
| Student / On a government training programme (Nation Traineeship/Modern Apprenticeship) | 4 |
| Out of work (6 months or less) | 5 |
| Out of work (more than 6 months) | 6 |
| Looking after home / Homemaker | 7 |
| Retired | 8 |
| Disabled OR Long-term sick | 9 |
| Unpaid work for a business, community or voluntary organisation | 10 |
| Prefer not to say | 11 |

(Code 1 if selected 1, 2 if selected 2 and 3 if selected 3. Others set to missing)

### 11. Occupational group

D4. **Which of the following best describes the occupation of the member of your household with the largest income (the chief income earner)? Please select one answer**

Please indicate to which occupational group the Chief Income Earner in your household belongs, or which group fits best.

The Chief Income Earner is the person in your household with the largest income.

If the Chief Income Earner is retired and has an occupational pension please answer for their most recent occupation.

If the Chief Income Earner is not in paid employment but has been out of work for less than 6 months, please answer for their most recent occupation
SINGLE CODE

| Semi or unskilled manual work (e.g. Manual workers, all apprentices to skilled trades, Caretaker, Park keeper, non-HGV driver, shop assistant) | 1 |
| --- | --- |
| Skilled manual worker *(e.g. Skilled Bricklayer, Carpenter, Plumber, Painter, Bus/ Ambulance Driver, HGV driver, AA patrolman, pub/bar worker, etc.)* | 2 |
| Supervisory or clerical/ junior managerial/ professional/administrative *(e.g. Office worker, Student Doctor, Foreman with 25+ employees, salesperson, etc.)* | 3 |
| Intermediate managerial/ professional/ administrative *(e.g. Newly qualified (under 3 years) doctor, Solicitor, Board director small organisation, middle manager in large organisation, principle officer in civil service/local government)* | 4 |
| Higher managerial/ professional/ administrative *(e.g. Established doctor, Solicitor, Board Director in a large organisation (200+ employees, top level civil servant/public service employee)* | 5 |
| Student | 6 |
| Casual worker – not in permanent employment | 7 |
| Housewife/ Homemaker | 8 |
| Retired and living on state pension | 9 |
| Unemployed or not working due to long-term sickness | 10 |
| Full-time carer of another household member | 11 |
| Other | 95 |

(Code 1 if selected 3 to 5 and 2 if selected 1 or 2)

### 12. Work sector

ASK IF D2= CODES 1,2, 3, 4, or 10

D2B. **Please could you indicate if you work in any of the following sectors or roles? Please include any voluntary work**

| Health or social care *(e.g. doctors, nurses, midwives, paramedics, social workers, care workers; or work as part of the health and social care supply chain, including producers and distributers of medicines and medical equipment)* | 1 |
| --- | --- |
| Education and Childcare *(e.g. teaching and support staff, childminders, social workers, specialist education professionals, etc.* | 2 |
| Key public services *(e.g. the justice system, religious staff, charities delivering frontline services, journalists, broadcasters, undertakers, etc.)* | 3 |
| Local and national government *(e.g. occupations essential to continuous provision of essential services, such as the payment of benefits, or processing of new benefit applications)* | 4 |
| Food and essential goods *(e.g. food production, processing, distribution, sale and delivery, as well as those essential to the provision of other key goods such as hygienic or veterinary medicine)* | 5 |
| Public safety and national security *(e.g. police and support staff, Ministry of Defence civilians, contractor and armed forces, fire and rescue service employees, National Crime Agency staff, border security staff, prison and probation staff and other national security roles)* | 6 |
| Transport *(e.g. air, water, road and rail passenger and freight transport modes)* | 7 |
| Utilities, communication and financial services *(e.g. banks, building societies and financial market infrastructure; the oil, gas, electricity and water sectors; information technology and data infrastructure sector; civil nuclear, chemicals, telecommunications, network operations, field engineering, call centre staff, IT and data infrastructure, 999 and 111 critical services, postal services and delivery, payments providers and waste disposal)* | 8 |
| None of the above | 95 |

### 13.  Index of Multiple Deprivation

Based on postcode as with Region using lookup table as given in reference.

### 14.  Financial hardship

GRID QUESTION. SINGLE CODE PER STATEMENT

RANDOMISE STATEMENTS

**Q24. Thinking now about the past seven days, could you tell us to what extent you agree or disagree with the following statements?**

SCALE:

1. Strongly agree
2. Agree
3. Neither agree nor disagree
4. Disagree
5. Strongly disagree
6. Not applicable

STATEMENTS:

1. I am finding my current living situation difficult
2. I am struggling to make ends meet
3. I am skipping meals I would usually have

(Reverse code 5 to 1 from strongly agree to strongly disagree and add scores to sum from 3 to 15)

### 15.  Chronic illness of self

D5. **Do you, or anyone else in your household have any long-standing illness, disability or infirmity?**

***Please select all that apply***

MULTI CODE

| 1 | **Yes, I do** |
| --- | --- |
| 2 | **Yes, another household member** |
| 3 | **No [EXCLUSIVE]** |
| 97 | **Prefer not to say [EXCLUSIVE]** |

IF D5=1
D6. **Do you have any of the following health conditions?**

***Please select all that apply***

MULTI CODE

| 1 | Diabetes (type 1) |
| --- | --- |
| 2 | Diabetes (type 2) |
| 3 | Epilepsy |
| 4 | Any type of cancer |
| 5 | Chronic Obstructive Pulmonary Disease (COPD), including emphysema and chronic bronchitis |
| 6 | Asthma |
| 7 | Chronic pain |
| 8 | Mental health conditions |
| 9 | Heart conditions |
| 17 | Chronic kidney disease |
| 10 | Chronic liver disease and cirrhosis |
| 18 | A condition affecting your brain and nerves, such as Parkinson's disease, motor neurone disease, multiple sclerosis (MS), a learning disability or cerebral palsy |
| 11 | Recovering from a stroke |
| 12 | A condition that makes you much more likely to get infections (e.g. SCID, homozygous sickle cell) |
| 13 | Have had an organ transplant |
| 14 | Have had a bone marrow or stem cell transplant in the last 6 months |
| 19 | Are taking medicine that weakens your immune system (e.g. steroid tablets, chemotherapy, or antiretroviral medications) |
| 20 | HIV/AIDS |
| 17 | Very overweight |
| 95 | Other |
| 97 | Prefer not to say [EXCLUSIVE] |

(Code 1 if D5=1 and D6=1 to 17)

### 16.  Chronic illness of household member

As with Chronic illness of self but with D5=2.

### 17. Vaccination status

**QVac1a. Have you received a coronavirus vaccine?**

1. Yes
2. No – The NHS has offered me the vaccine but I have not had it
3. No - I have not received a coronavirus vaccine nor been invited to have one by the NHS
4. Don’t know

(Code 1 if selected 1 and 0 otherwise)

### 18.  Worried about Covid

SINGLE CODE

**Overall, how worried are you about coronavirus?**

1. Extremely worried
2. Very worried
3. Somewhat worried
4. Not very worried
5. Not at all worried
6. Don’t know

(Reverse code 5 to 1 with 6 set to missing)

### 19.  Risk of Covid to self

**To what extent do you think coronavirus poses a risk to:**

SCALE:

1. Major risk
2. Significant risk
3. Moderate risk
4. Minor risk
5. No risk at all
6. Don’t know

STATEMENTS:

- To you personally?

(Reverse code with 5 set to 1 with 6 set to missing)

### 20.  Believe Government information is biased

SINGLE CODE

**To what extent do you agree or disagree with the following statements:**

SCALE:

1. Strongly agree
2. Agree
3. Neither agree nor disagree
4. Disagree
5. Disagree strongly
6. Don’t know

STATEMENTS:

- Information from the Government about coronavirus is biased or one-sided

(Reverse code with 5 set to 1 with 6 set to missing)

### 21.  Believe had Covid

**Q23. Do you know if you have had, or currently have, coronavirus?**

1. I’ve definitely had it, and had it confirmed by a test
2. I think I’ve probably had it
3. I don’t know whether I’ve had it or not
4. I think I’ve probably not had it
5. I’ve definitely not had it

(Code 1 or 2 to 1 and others to 0)

### 22.  Willing to leave home with symptoms

ROUTING: ASK IF DO NOT ANSWER ‘NEW, CONTINUOUS COUGH’ OR ‘HIGH TEMPERATURE’ OR ‘LOSS OF SENSE OF SMELL’ OR ‘LOSS OF TASTE’ FOR Q14

**Q17B. Again, imagine that tomorrow morning you develop symptoms of coronavirus (high temperature/fever, a new, continuous cough, or a loss of taste or smell).**

**Could you tell us what, if anything, would cause you to leave the home during this time?**

**Please select all that apply.**

1. If my symptoms were only mild
2. If my symptoms got better
3. If my symptoms got worse
4. If my symptoms did not persist / were temporary
5. For a medical need (other than coronavirus)
6. To go to the shops, for groceries/pharmacy
7. To go to the shops, for things other than groceries/pharmacy
8. To go for a walk or some other exercise
9. To go out to work
10. To help or provide care for a vulnerable person
11. To meet up with friends and/or family
12. I don’t think it is necessary for me to stay at home
13. If I became too depressed or anxious
14. If I became too lonely
15. If I became too bored
16. I would not leave the home at all [SINGLE CODE]
17. Other, please specify

(Code 1 if selected any of 1 to 15, 0 if 16)

### 23.  Nonessential workplace attendance

**D2E.** **If you wanted to, are you able to work from home?**

1. Yes – full time
2. Yes – most of the time
3. Yes – but I have to go into my place of work once or twice a week
4. No – I cannot do my job from home

GRID QUESTION. SINGLE CODE PER STATEMENT

**Q8_C. Please enter the number of times you have been out of your home in the last seven days, for each of the following reasons?**

**If you have not left your home for this reason, please write 0**

RANDOMISE ORDER, ANCHOR OTHER

STATEMENTS

1. To go to the shops, for groceries/pharmacy
2. To go to the shops, for things other than groceries/pharmacy
3. To go for a walk or some other exercise
4. To spend time outdoors for recreational purposes (including to sit in parks etc.)
5. To go out to work
6. To take a child (or children) to/from school
7. For a medical need, or to donate blood
8. To travel to a different area (including for day trips)
9. To help or provide care for a vulnerable person
10. To meet up with friends and/or family that you don’t live with
11. To go to a restaurant, café or pub
12. To go to a place of worship
13. Other, please specify

(Code 1 if selected 1 to D2E and entered number greater than 0 to Q8_C)
